# COVID-19 Mortality in California Based on Death Certificates: Disproportionate Impacts Across Racial/Ethnic Groups and Nativity

**DOI:** 10.1101/2021.03.01.21252678

**Authors:** Erika Garcia, Sandrah P. Eckel, Zhanghua Chen, Kenan Li, Frank D. Gilliland

**Author notes:** Corresponding Author Erika Garcia, Keck School of Medicine of USC, Department of Preventive Medicine, 2001 N. Soto Street, SSB1 Rm 225F MC 9237, Los Angeles, CA 90089-9237.

## Abstract

**Purpose:** To examine characteristics of coronavirus disease 2019 (COVID-19) decedents in California (CA) and evaluate for disproportionate mortality across race/ethnicity and ethnicity/nativity.

**Methods:** COVID-19 deaths were identified from death certificates. Age-adjusted mortality rate ratios (MRR) were compared across race/ethnicity. Proportionate mortality rates (PMR) were compared across race/ethnicity and by ethnicity/nativity.

**Results:** We identified 10,200 COVID-19 deaths in CA occurring February 1 through July 31, 2020. Decedents tended to be older, male, Hispanic, foreign-born, and have lower educational attainment. MRR indicated elevated COVID-19 morality rates among Asian/Pacific Islander, Black, and Hispanic groups compared with the White group, with Black and Hispanic groups having the highest MRR at 2.75 (95%CI:2.54-2.97) and 4.18 (95%CI: 3.99-4.37), respectively. Disparities were larger at younger ages. Similar results were observed with PMR, which remained in analyses stratified by education. Elevated PMR were observed in all ethnicity/nativity groups, especially foreign-born Hispanic individuals, relative to U.S.-born non-Hispanic individuals, were generally larger at younger ages, and persisted after stratifying by education.

**Conclusions:** Differential COVID-19 mortality was observed in California across racial/ethnic groups and by ethnicity/nativity groups with evidence of greater disparities among younger age groups. Identifying COVID-19 disparities is an initial step towards mitigating disease impacts in vulnerable communities.

## INTRODUCTION

The is a growing body of literature on the differential impacts of the coronavirus disease 2019 (COVID-19) in historically marginalized groups in the United States (U.S.), including disease incidence,^1–3^ hospitalization,^4–8^ severity,^8–10^ and mortality.^2,6,8,11–13^ Identifying COVID-19 disparities is an initial step towards mitigating disease impacts in vulnerable communities. Much of the current evidence, however, has relied on ecologic analyses using aggregate data for both the characteristic under study (e.g., percent Black in a county) and COVID-19 outcome (e.g., county-level mortality).^1,2,11,13^ Studies that have been conducted with individual-level data primarily derive from hospital/healthcare networks or insurance companies,^3–6,9,14^ which may not be representative of COVID-19 impacts in the general population given barriers to healthcare access and insurance among historically marginalized groups. Use of death certificate data allows for investigation into individual-level characteristics and captures all individuals in the catchment area. Appropriate estimates of disease are needed, not only by single factors, such as racial/ethnic group, but also by intersections with other factors, such as with age, sex, and educational attainment, which considers that groups are not homogenous and indeed reflect diverse experiences and impacts of the pandemic.^15–17^ Intersectional analysis allows for a more nuanced understanding of disparate burden of COVID-19, which is crucially needed to design and implement effective public health interventions to mitigate disease impacts.

The objective of this study was to examine demographic characteristics of COVID-19 decedents in California (CA)—which as of September 2020 is amongst the hardest hit states in terms of cases and deaths^18^—and evaluate for disproportionate mortality across these characteristics. In addition to descriptive information on decedents, COVID-19 mortality was compared across racial/ethnic groups considering age, sex, and education using mortality rate ratios (MRR) and proportionate mortality ratios (PMR). We also examined COVID-19 mortality by ethnicity and nativity combined, considering age and education, with PMR.

## METHODS

### Study Setting and Population

Data on causes of death, race/ethnicity, sex, age, educational attainment, country of birth, and county of residence were obtained using the California Comprehensive Death Files (CCDF) and California Comprehensive Master Death File (CCMDF) from the California Department of Public Health, Center for Health Statistics and Informatics for 2016 to 2020 (CCMDF for 2016-2018 and CCDF for 2019-2020). Data for 2020 were updated weekly and last date of data export used in this analysis was September 9, 2020. Race/ethnicity were grouped into non-Hispanic White, non-Hispanic Black, Asian/Pacific Islander (regardless of Hispanicity; Asian/PI), American Indian/Alaskan Native (regardless of Hispanicity; AI/AN), Hispanic (excludes A/PI, AI/AN), and Multiracial/Other/Unknown. County of birth was used to define decedents ‘nativity status, categorized as U.S.-born or foreign-born. Categories of ethnicity and nativity combined were U.S.-born non-Hispanic, U.S.-born Hispanic, Foreign-born non-Hispanic, and Foreign-born Hispanic, using Hispanicity as defined for race/ethnicity. This study was approved by the Committee for the Protection of Human Subjects of the state of CA.

### COVID-19-Related Deaths

Mortality data included the 10th revision of the *International Statistical Classification of Diseases and Related Health Problems* (ICD-10) codes for underlying cause of death and up to 20 relevant conditions. Effective April 1, 2020, there was a new ICD-10 code for COVID-19, U07.1; however, because we used the dynamic CCDF file with weekly updates for recent mortality, final coded data on underlying cause of death and relevant conditions were not available for all 2020 deaths. Thus, we developed an algorithm using the code U07.1 and a keyword search to identify COVID-19-related deaths and applied it to all deaths occurring after February 1, 2020 (eFigure 1). Among deaths with coded data on underlying cause of death and relevant conditions, our algorithm corrected identified 99.95% of deaths coded with U07.1. Analyses were restricted to COVID-19-related deaths occurring February 1-July 31, 2020.

### Statistical Analysis

Temporal trends in COVID-19 mortality in CA during the study period are presented. Descriptive statistics among decedents identified as have COVID-19 mortality were presented for the entire study period as well as for sub-periods with distinct patterns in mortality (i.e., distinct “epidemic periods”).

COVID-19 age-adjusted mortality rates (MR) per 100,000 person-years were calculated using direct standardization to the 2019 CA population using 5-year age intervals and were compared across racial/ethnic groups using mortality rate ratios (MRR) by sex and epidemic period, accounting for variation in observation time. Non-Hispanic White was used as the referent group. Population estimates were obtained from the CDC WONDER Online Database using the Bridged-Race Population Estimates for CA^19^ and racial/ethnic groups were combined to align with categories described above (does not include “Multiracial/Other/Unknown” group). Age-adjusted MR were also calculated and compared across racial/ethnic groups using MRR for selected counties with >500 COVID-19 deaths using 2019 county-specific population estimates for both population and direct standardization. COVID-19 age-specific MR were calculated and compared across racial/ethnic groups using MRR by age groups (excluding 0-19 years category, which had only 5 deaths) and sex.

Analyses using proportionate mortality (PM) were restricted to deaths among decedents aged 20 years and older and occurring March 1-July 31 (only 2 COVID-19 deaths occurred before March 1, 2020). PM was defined as number of COVID-19 deaths in a specified subpopulation (e.g., a certain racial/ethnic group) divided by average all-cause mortality in the same sub-population occurring March 1-July 31 in 2016-2019 among decedents aged 20 years and older (multiplied by 100 to be akin to a percentage). The PM metric allows for COVID-19 mortality to be interpreted as a percentage of “typical” mortality in a group. Mortality for past years was restricted to match the time frame for 2020 COVID-19 deaths to account for annual trends in mortality. All-cause mortality in 2020 was not used as the reference, which would have been common for PM, because evidence suggests all-cause mortality has been affected by the pandemic not only through COVID-19 mortality.^20^ PM ratios (PMR) were used to compare across decedent characteristics, including racial/ethnic group, sex, education attainment, and ethnicity and nativity combined, as well as combinations of these. Educational attainment may serve as a proxy for socioeconomic status which is linked with healthcare access, co-morbidities, and other factors such as employment type that may impact COVID-19 infection and mortality.^21–24^ Because the PMR metric relies on all-cause mortality, we additionally examined differences in all-cause mortality rates for 2019 by age among racial/ethnic groups using 2019 death data and population estimates.

All analyses were performed using SAS statistical software version 9.4 (SAS Institute Inc., Cary, NC) and statistical tests were based on 2-sided tests with α=0.05. Standard errors and 95% confidence intervals (95%CI) were computed using standard methods (see eMethods).^25^

## RESULTS

We identified 10,200 COVID-19 deaths occurring in CA between February 1 and July 31, 2020. Distinct temporal patterns in weekly mortality identified three periods in the COVID-19 epidemic in CA (eFigure 2): the first period, through April 19, had an upward trend; the second period, April 20-June 16, demonstrated a steady slight decline; the third period, June 17-July 31, had an upward trend similar to the first period. Distribution of characteristics among decedents with COVID-19 mortality are shown for the entire study period (Table 1) and by the three epidemic periods (eTable 1). Decedents were consistently older, more likely to be male, had lower educational attainment, and most were foreign-born. Hispanic was the predominant race/ethnicity among decedents, which increased proportionately during the epidemic.

**Table 1.** **Distribution of selected characteristics among decedents identified as having COVID-19 mortality**.

|  |  |
| --- | --- |
|  | **N (%)** |
| Deaths | 10200 |
| **Age** |  |
| 0 to 19 | 5 (0.1) |
| 20 to 44 | 421 (4.1) |
| 45 to 54 | 710 (7.0) |
| 55 to 64 | 1467 (14.4) |
| 65 to 74 | 2222 (21.8) |
| 75 to 84 | 2450 (24.0) |
| 85 or older | 2925 (28.7) |
| **Sex** |  |
| Male | 5914 (58.0) |
| Female | 4286 (42.0) |
| **Race/ethnicity** |  |
| Hispanic (Excludes A/PI, AI/AN) | 4914 (48.2) |
| *Mexican* | 3333 (67.8) |
| *Other Hispanic* | 1581 (32.2) |
| NH White | 3025 (29.7) |
| Asian/Pacific Islander | 1290 (12.7) |
| *Filipino* | 441 (34.2) |
| *Chinese* | 251 (19.5) |
| *Korean* | 207 (16.1) |
| *Vietnamese* | 97 (7.5) |
| *Japanese* | 82 (6.4) |
| *Other Asian* | 212 (16.4) |
| Black | 804 (7.9) |
| Multiracial/Other/Unknown | 124 (1.2) |
| American Indian/Alaskan Native | 43 (0.4) |
| **Educational Attainment** |  |
| Less than High School | 3637 (35.7) |
| High School | 3027 (29.7) |
| Some college | 1040 (10.2) |
| Associate Degree | 478 (4.7) |
| Bachelor's Degree | 966 (9.5) |
| Graduate Degree | 499 (4.9) |
| Unknown | 553 (5.4) |
| **Country of Birth** |  |
| United States | 4413 (43.3) |
| Mexico | 2942 (28.8) |
| Philippines | 442 (4.3) |
| El Salvador | 364 (3.6) |
| Guatemala | 219 (2.2) |
| South Korea | 195 (1.9) |
| Other country | 1302 (12.8) |
| Unknown | 323 (3.2) |

### Mortality Rate Ratios

Morality rates were considerably higher among older individuals but with differences observed across racial/ethnic groups (Figure 1). Compared with the White group, MRR were elevated among Asian/PI, Black, and Hispanic groups, with Black and Hispanic groups having the highest MRR at 2.75 (95%CI:2.54-2.97) and 4.18 (95%CI:3.99-4.37), respectively (Figures 2 and eFigure 3; eTable 2). This pattern persisted within each epidemic period (eFigure 3). Analyses by county of residence for counties with >500 COVID-19 deaths (Los Angeles, Orange, Riverside, San Bernardino, and San Diego Counties) showed that while there were some differences in specific MRRs estimated by racial/ethnic group, the overall pattern of COVID-19 mortality disparity persisted (eFigure 4).

**Figure 1.**
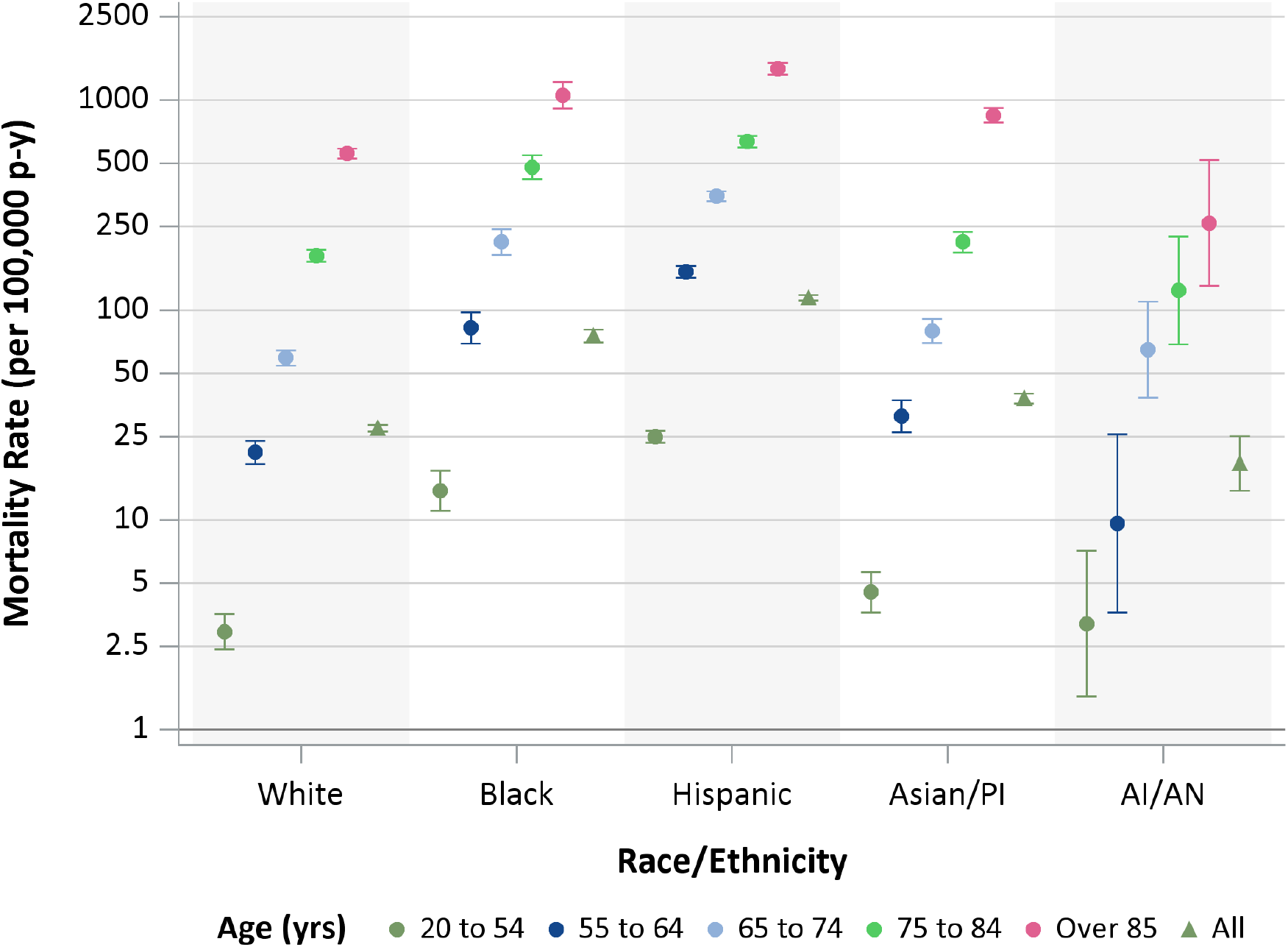
Age-standardized COVID-19 mortality rates for all ages and select ages by race/ethnicity. Mortality rates are per 100,000 person-years. Bars represent 95% confidence intervals.

**Figure 2.**
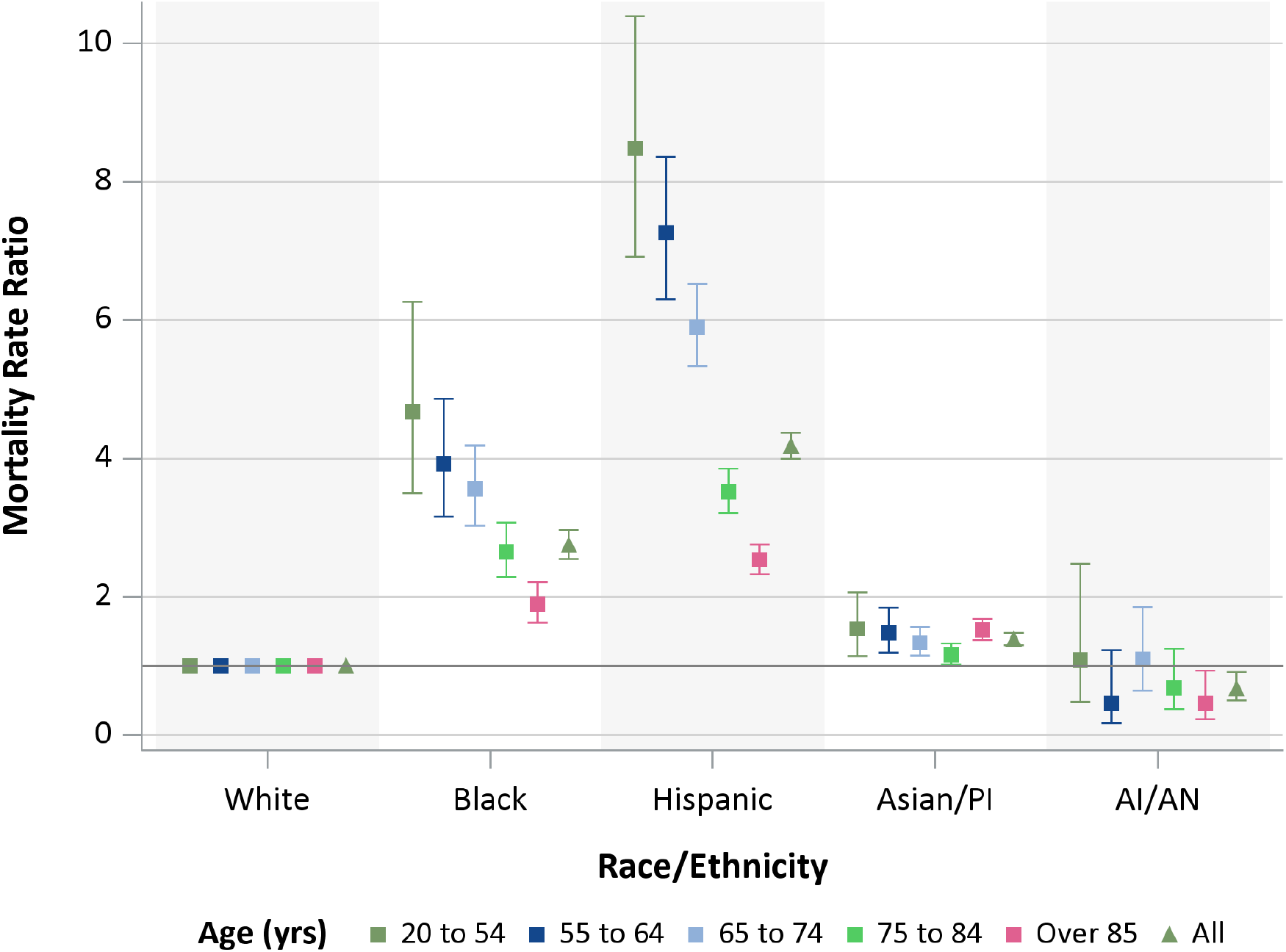
Age-standardized COVID-19 mortality rate ratios for all ages and select ages by race/ethnicity. Bars represent 95% confidence intervals. Referent group is non-Hispanic Whites.

Age-specific MRR revealed greater differences among racial/ethnic groups, with greater disproportionate mortality among younger Black and Hispanic individuals (Figure 2; ETable 3). Compared with Whites individuals aged 20-54 years, the MR was 4.7-fold (95%CI:3.5-6.3) higher among Black individuals and 8.5-fold (95%CI:6.9-10.4) higher among Hispanic individuals of the same age. When examined by sex, disparities observed among decedents aged less than 84 years were higher among females in the Black group and among males in the Hispanic group. In the Asian/PI group, mortality disparity was greatest among those aged 85 year or more, with the largest MRR observed for female, 1.70 (95%CI:1.50-1.93).

### Proportionate Mortality

Univariate analyses using PMR found elevated PM in males compared with females, in all non-White racial/ethnic groups, in decedents with lower education attainment, and in all ethnicity/nativity groups relative to U.S.-born non-Hispanic (eTable 4).

Analyses comparing across racial/ethnic groups by age demonstrated greater disparities among younger decedents, similar to those observed with MRR (eTable 5). Elevated PMR persisted in analyses stratified by education (Figure 3; eTable 6). For example, among decedents aged 20-64 years with high school education or less, the PM in the Hispanic group was 8.5-fold (95%CI:7.4-9.9) higher compared with the White group. For the same age group, but among those with some college or more the PMR for Hispanics remained highly elevated at 7.0 (95%CI:5.8-8.6). All-cause mortality rates for 2019 were lower compared with the White group for all racial/ethnic group, except the Black group which were higher. This may lead PMR to over or underestimate true COVID-19 mortality disparities which should be considered when interpreting these results, but these differences were not sufficiently large to explain away all disparities observed (eFigure 5). Adjusting PMR for difference in all-cause mortality produces estimates similar to MRR reported above (eTable 7). Examining ethnicity and nativity combined by age, elevated PMR were observed for all groups, especially for the foreign-born Hispanic group, relative to the U.S.-born non-Hispanic group and PMR were generally larger among younger age groups (eTable 8). After accounting for educational attainment by stratification, elevated PMR remained (Table 2). Among decedents aged 20-64 years, PMR for foreign-born Hispanic compared with U.S.-born non-Hispanic among those with a high school education or less was PMR=10.7 (95%CI:9.5-12.1) while among those with some college or more was PMR=8.4 (95%CI:6.9-10.1).

**Figure 3.**
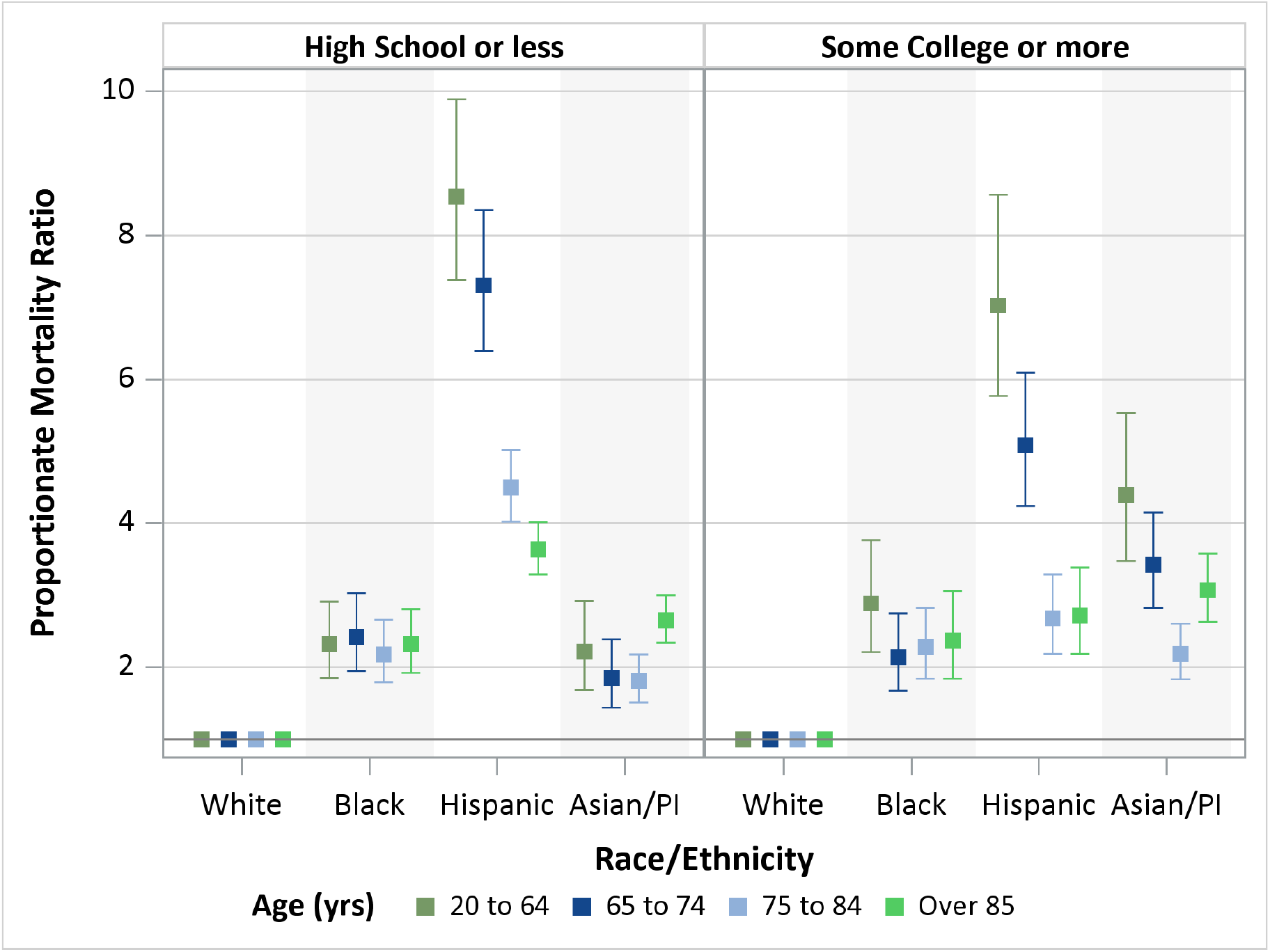
Proportionate mortality ratios for COVID-19 by race/ethnicity, age, and educational attainment. Proportionate mortality ratios are based on COVID-19 deaths occurring between March 1-July 31, 2020 among decedents aged 20 years and older. Sub-population proportionate mortalities were calculated using all-cause mortality in the same sub-population occurring March 1-July 31 in 2016-2019 among decedents aged 20 years and older. Bars represent 95% confidence intervals. Referent group for PMR is non-Hispanic White.

**Table 2.** **Proportionate mortality (PM) and proportionate mortality ratio (PMR) by ethnicity and nativity combined, age, and educational attainment for COVID-19 deaths occurring between March 1 - July 31, 2020 among decedents aged 20 years and older**.

| Ethnicity and Nativity Combined | Age | High school or less |  |  | Some college or more |  |  |
| --- | --- | --- | --- | --- | --- | --- | --- |
|  |  | COVID-19 Deaths | PM (95% CI)<sup>a</sup> | PMR (95% CI) | COVID-19 Deaths | PM (95% CI)<sup>a</sup> | PMR (95% CI) |
| U.S.-born non-Hispanic | 20 to 64 | 295 | 3.6 (3.2-4.0) | Referent | 223 | 3.1 (2.7-3.5) | Referent |
|  | 65 to 74 | 309 | 5.7 (5.1-6.3) | Referent | 315 | 4.3 (3.8-4.8) | Referent |
|  | 75 to 84 | 441 | 6.1 (5.6-6.7) | Referent | 468 | 5.4 (4.9-5.9) | Referent |
|  | 85<= | 602 | 4.9 (4.6-5.3) | Referent | 532 | 4.4 (4.0-4.8) | Referent |
| U.S.-born Hispanic | 20 to 64 | 272 | 10.1 (9.0-11.4) | 2.81 (2.40-3.29) | 130 | 10.4 (8.8-12.4) | 3.40 (2.76-4.18) |
|  | 65 to 74 | 170 | 18.1 (15.6-21.1) | 3.20 (2.69-3.80) | 67 | 13.0 (10.2-16.5) | 3.06 (2.39-3.92) |
|  | 75 to 84 | 166 | 14.0 (12.0-16.3) | 2.29 (1.94-2.71) | 40 | 9.2 (6.8-12.6) | 1.71 (1.26-2.33) |
|  | 85<= | 236 | 12.1 (10.7-13.8) | 2.46 (2.13-2.84) | 33 | 8.6 (6.1-12.1) | 1.96 (1.40-2.74) |
| Foreign-born non-Hispanic | 20 to 64 | 84 | 6.7 (5.4-8.4) | 1.87 (1.48-2.37) | 138 | 7.9 (6.7-9.3) | 2.57 (2.09-3.16) |
|  | 65 to 74 | 109 | 9.3 (7.7-11.2) | 1.63 (1.32-2.01) | 180 | 11.5 (10.0-13.4) | 2.71 (2.27-3.23) |
|  | 75 to 84 | 224 | 10.0 (8.8-11.4) | 1.63 (1.40-1.90) | 207 | 9.5 (8.3-10.9) | 1.77 (1.51-2.07) |
|  | 85<= | 506 | 12.1 (11.1-13.2) | 2.46 (2.20-2.75) | 265 | 10.1 (8.9-11.4) | 2.30 (1.99-2.64) |
| Foreign-born Hispanic | 20 to 64 | 1177 | 38.7 (36.6-41.0) | 10.73 (9.51-12.11) | 161 | 25.6 (21.9-29.9) | 8.35 (6.93-10.05) |
|  | 65 to 74 | 792 | 45.2 (42.1-48.4) | 7.96 (7.06-8.98) | 94 | 31.5 (25.7-38.5) | 7.39 (6.06-9.02) |
|  | 75 to 84 | 691 | 32.4 (30.1-35.0) | 5.31 (4.76-5.92) | 59 | 19.2 (14.9-24.7) | 3.55 (2.78-4.54) |
|  | 85<= | 547 | 23.7 (21.8-25.8) | 4.80 (4.31-5.34) | 50 | 15.2 (11.5-20.0) | 3.46 (2.64-4.52) |

<sup>a</sup> Sub-population PM were calculated using all-cause mortality in the same sub-population occurring March 1-July 31 in 2016-2019 among decedents aged 20 years and older.

## DISCUSSION

This study used death certificate data to identify decedents with COVID-19 mortality in CA and investigate demographic characteristics associated with mortality using two different metrics of association, MRR and PMR. Disproportionate COVID-19 mortality was observed among Black, Hispanic, and Asian/PI groups in CA. Larger relative disparities were observed at younger ages for Black and Hispanic individuals. Disparities persisted after accounting for educational attainment, serving here as a proxy for socioeconomic status. Disproportionate mortality was also observed by combinations of ethnicity and nativity, with all groups, but especially foreign-born Hispanic individuals, having greater COVID-19 mortality compared with U.S.-born non-Hispanic individuals, particularly at younger ages. Disparities by nativity remained in analyses controlling for educational attainment. The larger disparities in younger age groups are particularly important given younger populations may be overlooked in public health campaign due to their overall lower risk of sever disease. Here, however, we observed mortality rate for Black individuals aged 55-64 to be higher than mortality rate for White individuals 10 years older, and mortality rate for Hispanic individuals aged 55-64 years to be approaching mortality rate for White individuals 20 years older. Younger populations should be included among targets in public health interventions. Disparities in COVID-19 mortality may be driven by a variety of factors, including complex interaction between social and structural determinates of health, barriers to accessing care, higher prevalence of underlying co-morbidities associated with more sever COVID-19 disease and adverse outcomes, and differential exposure to virus due to working and living conditions.^21,26–30^

Differences in COVID-19 mortality can develop anywhere along the disease pathway from exposure, incidence, severity, through deaths, and may different for different groups (e.g., younger Black group, older Asian/PI, etc.). Several studies have reported differential COVID-19 impacts across racial/ethnic groups, although few studies have examined outcomes by nativity. An ecologic study in Massachusetts found higher population percent Black and Latino to positively associate with COVID-19 incidence rates. Percent foreign-born non-citizens living in a community, mean household size, and share of food service workers were also positively associated with incidence, and after accounting for these factors the association for the Latino population with incidence was attenuated, but for the Black population was not.^31^ COVID-19 hot spots in New York City and Chicago tended to be in areas with higher proportions of Black residents, and in New York City in area with higher proportions of Hispanic and foreign-born residents.^23^ In a separate study across 22 states, disproportionate COVID-19 incidence was reported for Hispanic, Black, AI/AN, Asian, and Native Hawaiian/Pacific Islander in counties considered hotspots for COVID-19 incidence.^1^ A healthcare system-based study reported non-Hispanic Black and Hispanic individuals as having higher odds of infection, adjusted for demographic and socioeconomic characteristics.^32^ Another healthcare system-based study using prefer spoken language as a surrogate for immigrant status found that compared with English-speakers, non-English speakers were less likely to have completed severe acute respiratory syndrome coronavirus 2 testing and that the proportion of positive cases was 4.6-fold higher.^33^ Higher hospitalization has been reported for Black and Hispanic patients, even after adjusting for co-morbidities and demographic and socioeconomic factors.^4,6,7^ Among patients hospitalized with COVID-19 infection, non-White patients were more likely to present with higher disease severity on admission chest radiographs, and increased severity was associated with worse outcomes.^10^ A study using aggregate data on COVID-19 from regions across the U.S. report disproportionately higher COVID-19 mortality relative to population size for Black individuals and higher estimated case-fatality.^12^ Among rural counties, average daily increase in COVID-19 mortality rates were significantly higher in counties with largest shares of Black and Hispanic residents.^13^ A study using electronic medical record data from 24 healthcare organization reported Black individuals having a greater odds of COVID-19 mortality, even after controlling for age, sex, and several co-morbidities.^24^ Differences between Black and White individuals were larger in those less than 50 years of age, similar to larger MRR and PMR observed in younger age groups in the present study. Not all studies examining race/ethnicity have observed difference in COVID-19 mortality. A large study using data from 92 U.S. hospitals found that after adjusting for a variety of factors including comorbidities, insurance, and neighborhood deprivation there was no difference in mortality between Black and White patients with COVID-19.^5^ Similarly, a study in Louisiana found that after adjusting for sociodemographic and clinical factors, in-hospital mortality was not different between Black and White patients.^6^ It may be that once COVID-19 patients are ill to the point of hospitalization, differences in mortality are less appreciable. Overall, this evidence taken together supports the notion that mortality differences across racial/ethnic groups and by ethnicity/nativity reported in the present study are likely the cumulative effect across the entire pathway of disease. Quantification of COVID-19 mortality disparities is needed so effective public health interventions can be developed to mitigate disproportionate burden of the pandemic on vulnerable populations.^34^ Factors that may contribute to mortality disparities and could be targets for interventions include barriers to healthcare access including medical mistrust and insurance, living and working conditions, being essential/frontline workers, underlying health conditions including suboptimal disease management, and social and structural determinates of health.^21,26–30,35^ Interactions between structural, social, and individual factors that contribute to differential COVID-19 mortality, however, are complex and vary not only across these different historically marginalized groups (e.g., Black, Hispanic, and Asian/PI), but also within these groups (e.g., younger and older Hispanic individuals). Public health interventions and policies should consider the differing and complex risk structure across and within these groups. For example, differential workplace exposure may contribute to increased mortality among younger Black and Hispanic, including foreign-born Hispanic, individuals. People of color are more likely to be employed in essential industries and in occupations with more exposure to infections and close proximity to others, and Black workers in particular face an elevated risk for these factors.^27^ A study of workplace COVID-19 outbreaks in Utah found 73% of cases were in Hispanic or non-White workers, whereas Hispanic and non-White workers represent only 24% of the workforce in affected industries.^30^ Immigrant families faced additional complicating factors which must be considered for public health planning. For example public charge regulations may disincentivize immigrant families from accessing healthcare if they become symptomatic, which delays testing and treatment and may increase disease transmission risk within their home and communities.^36,37^

This study has several strengths. First, to identify COVID-19 mortality we used death certificate data which captures all mortality in the state and does not restrict to individuals in a healthcare/insurance network. Second, we had information on individual-level demographic characteristics on decedent rather than relying on aggregate demographic data which could be subject to the ecologic fallacy. Third, because of this individual-level data we were able to examine mortality by several demographic factors in combination to better understand intersectional impacts of COVID-19. This study also has limitations. First, while use of death certificate data has many advantages, there are a limited number of available demographic and few socioeconomic variables. Augmentation of these data with neighborhood characteristics based on decedent residential address is an area of future work. Second, PM, and thus PMR, uses as a comparison all-cause mortality, which if different between groups may over or underestimate differences in mortality between groups. When all-cause mortality rates are lower compared with the referent group, PMR will overestimate differences in COVID-19 mortality but as discussed differences in all-cause mortality rates were not sufficient to explain away all observed COVID-19 disparities. When all-cause mortality rates are higher, however, such as was observed for Black individuals the PMR will underestimate differences COVID-19 mortality.

## CONCLUSIONS

Differential COVID-19 mortality was observed in California across racial/ethnic groups and by ethnicity and nativity combined, with evidence of greater disparities among younger age groups. Patterns of disparities persisted after accounting for educational attainment. Drivers of these disparate COVID-19 impacts of are likely multi-factorial and involve the interplay of structural, social, and individual factors that must be considered in the design and implementation for public health interventions to effectively mitigate the impacts of this disease.

## Supporting information

eTable

## Data Availability

The data used in this analysis are not available for replication because specific approvals from the Committee for the Protection of Human Subjects of the state of California must be obtained in order to access them.

## LIST OF ABBREVIATIONS AND ACRONYMS

AI/AN: American Indian/Alaskan Native
A/PI: Asian/Pacific Islander
CA: California
CCDF: California Comprehensive Death Files
CCMDF: California Comprehensive Master Death File
COVID-19: Coronavirus disease 2019
ICD-10: International Statistical Classification of Diseases and Related Health Problems
MR: Mortality rates
MRR: Mortality rate ratios
PM: Proportionate mortality
PMR: Proportionate mortality rates
U.S.: United States

## ACKNOWLEDGEMENTS

This research was supported by the National Institute of Environmental Health Sciences (P30ES007048), the Hastings Foundation, and by the Keck School of Medicine of USC COVID-19 Research Fund through a generous gift from the W. M. Keck Foundation.

