## Supplementary material for "COVID-19 Mortality in California Based on Death Certificates: Disproportionate Impacts Across Racial/Ethnic Groups and Nativity": eTable

**eMethods.** Statistical Analysis.

**eFigure 1.** Flow chart depicting algorithm used to identify COVID-19-related deaths.

**eFigure 2.** Weekly COVID-19 mortality in California during the study period calculated as a moving average using a 7-day window.

**eTable 1.** Distribution of selected characteristics among decedents identified as having COVID-19 mortality by period of the epidemic in California, N (%).

**eFigure 3.** Age-standardized mortality rate ratios (MRR) and 95% CI by race/ethnicity and study period.

**eTable 2.** Age-standardized mortality rates (MR) and mortality rate ratios (MRR) by sex, race/ethnicity, and study period. MR are per 100,000 person-years.

**eFigure 4.** Age-standardized mortality rate ratios (MRR) and 95% CI by race/ethnicity for selected counties.

**eTable 3.** Age stratum-specific mortality rates (MR) and mortality rate ratios (MRR) by race/ethnicity and sex. MR are per 100,000 person-years.

**eTable 4.** Proportionate mortality (PM) and proportionate mortality ratio (PMR) by selected characteristics for COVID-19 deaths occurring between March 1 - July 31, 2020 among decedents aged 20 years and older.

**eTable 5.** Proportionate mortality (PM) and proportionate mortality ratio (PMR) by race/ethnicity and age for COVID-19 deaths occurring between March 1 - July 31, 2020 among decedents aged 20 years and older.

**eTable 6.** Proportionate mortality (PM) and proportionate mortality ratio (PMR) by race/ethnicity, age, and educational attainment for COVID-19 deaths occurring between March 1 - July 31, 2020 among decedents aged 20 years and older.

**eFigure 5.** Crude all-cause mortality rate ratios and 95% CI based on 2019 mortality data and population estimates by race/ethnicity and age.

**eTable 7.** Crude all-cause mortality rate ratios for 2019, proportionate mortality ratio (PMR) for COVID-19, and adjusted PMR by race/ethnicity and age. The PMR are for COVID-19 deaths occurring between March 1 - July 31, 2020 among decedents aged 20 years and older.

**eTable 8.** Proportionate mortality (PM) and proportionate mortality ratio (PMR) by ethnicity and nativity combined and age for COVID-19 deaths occurring between March 1 - July 31, 2020 among decedents aged 20 years and older.

This supplementary material has been provided by the authors to give readers additional information about their work.

**eMethods.** Statistical Analysis

The SE for each individual MR and PM was calculated as follows: $SE[\ln(MR)] = 1/A^{1/2}$, where $A$ is the number of COVID-19 deaths. The standard error for each individual MRR and PMR was calculated as follows: $SE[\ln(MRR)] = (1/A_1 + 1/A_0 - 1/N_1 - 1/N_0)^{1/2}$, where $A_1$ is the number of COVID-19 deaths, $A_0$ is the number of COVID-19 deaths in the referent group, $N_1$ is the population (or comparison deaths for PMR) and $N_1$ is the population for the referent group (or comparison deaths for the referent group for PMR). The 95% CI for each measure of disease or association was calculated as $95\%\ CI = measure\ \pm 1.96 \times SE$.

**eFigure 1. Flow chart depicting algorithm used to identify COVID-19-related deaths.**

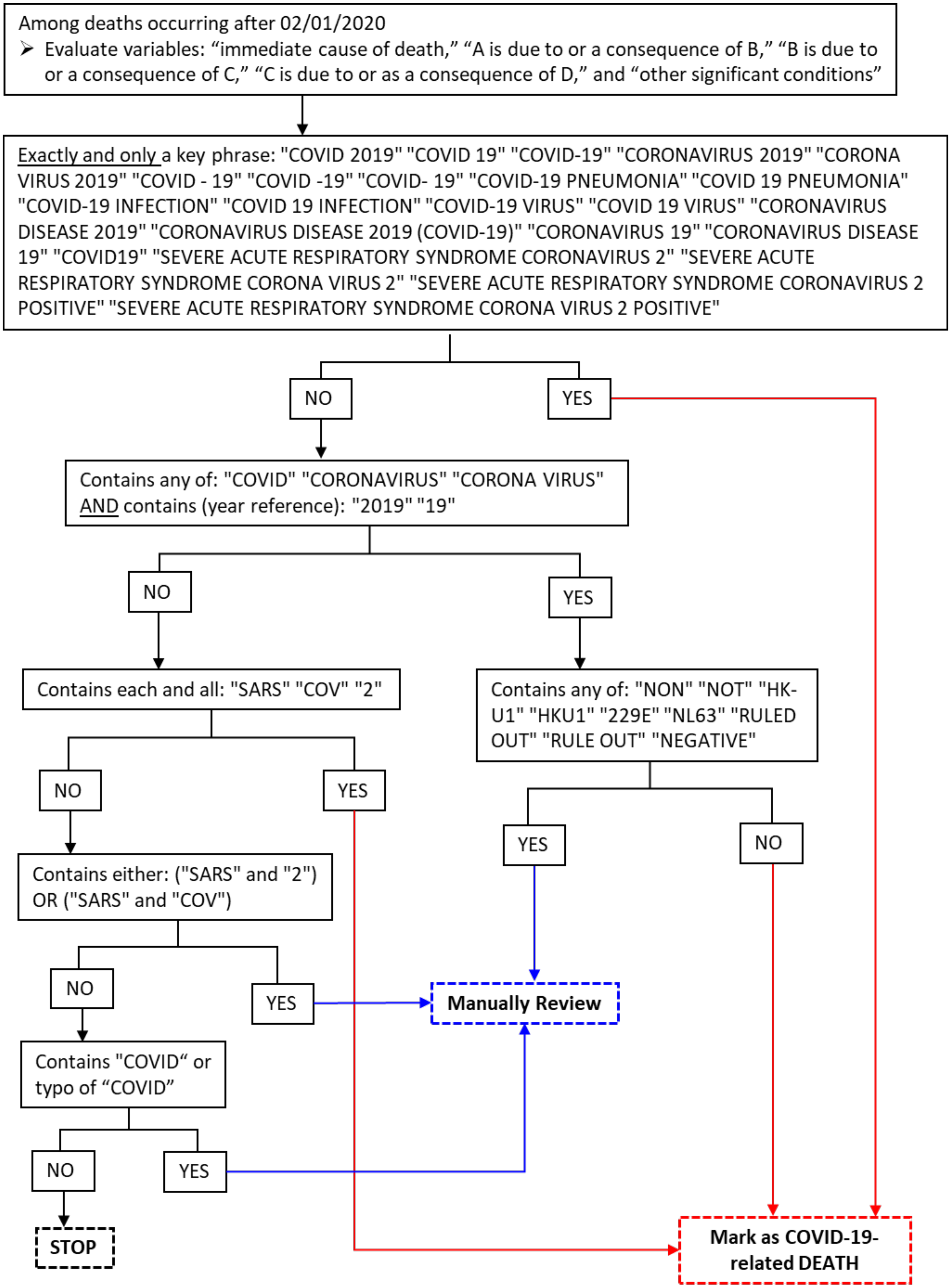


In addition to relying on the code U07.1 as the underlying cause of death or a relevant condition, we searched the following text fields for keywords or combinations of keywords: "immediate cause of death," "A is due to or a consequence of B," "B is due to or a consequence of C," "C is due to or as a consequence of D," and "other significant conditions". For select keywords or combinations of keywords, deaths were manually reviewed by the study team to evaluate if the death was COVID-19 related.

### eFigure 2. Weekly COVID-19 mortality in California during the study period calculated as a moving average using a 7-day window.

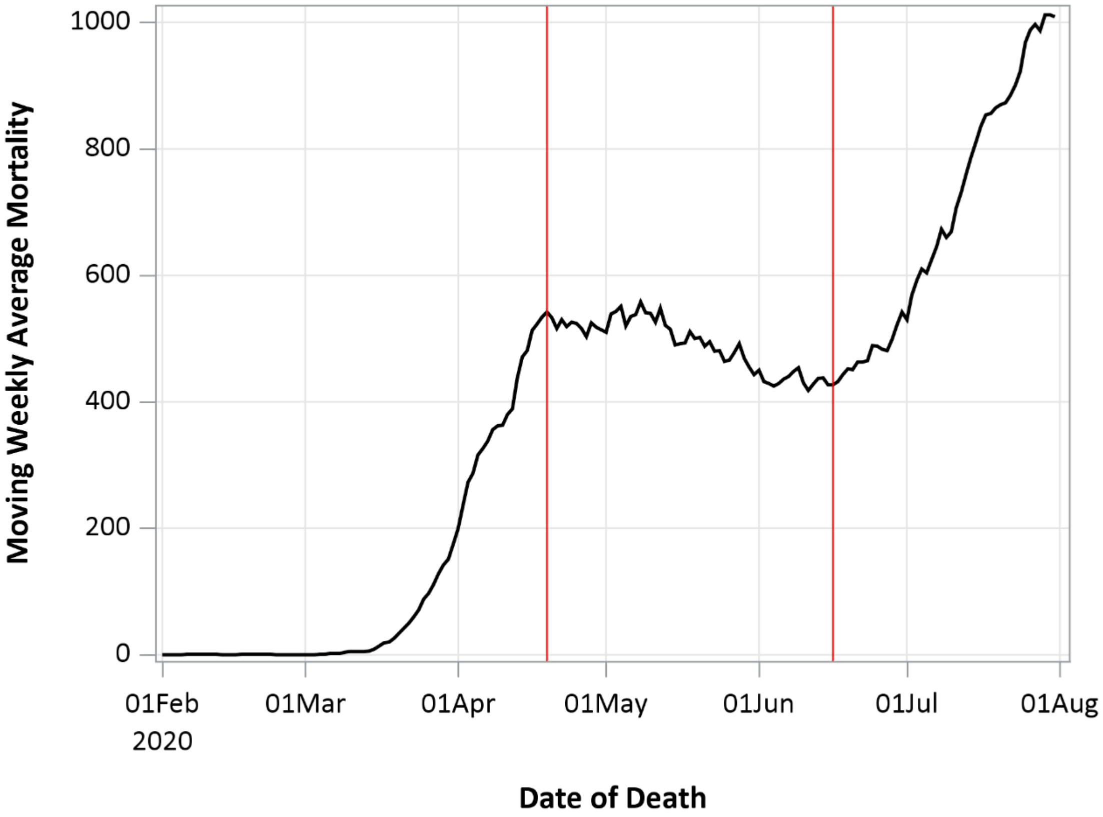


Red vertical lines separate the three periods of the epidemic in California with distinct patterns. Dates of separation are April 19 and June 16, 2020.

| **eTable 1. Distribution of selected characteristics among decedents identified as having COVID-19 mortality by period of the epidemic in California, N (%).** | | | |
|---|---|---|---|
| | **Period 1** | **Period 2** | **Period 3** |
| Date Range | 02/06/20-04/19/20 | 04/20/10-06/16/20 | 06/17/20-07/31/20 |
| Deaths | 1452 | 4017 | 4731 |
| **Age** | | | |
| 0 to 19 | 0 (0) | 1 (0) | 4 (0.1) |
| 20 to 44 | 66 (4.6) | 123 (3.1) | 232 (4.9) |
| 45 to 54 | 96 (6.6) | 249 (6.2) | 365 (7.7) |
| 55 to 64 | 175 (12.1) | 529 (13.2) | 763 (16.1) |
| 65 to 74 | 304 (20.9) | 838 (20.9) | 1080 (22.8) |
| 75 to 84 | 369 (25.4) | 986 (24.6) | 1095 (23.2) |
| 85 or older | 442 (30.4) | 1291 (32.1) | 1192 (25.2) |
| **Sex** | | | |
| Male | 861 (59.3) | 2264 (56.4) | 2789 (59.0) |
| Female | 591 (40.7) | 1753 (43.6) | 1942 (41.1) |
| **Race/ethnicity** | | | |
| Hispanic (Excludes A/PI, AI/AN) | 480 (33.1) | 1852 (46.1) | 2582 (54.6) |
| *Mexican* | *300 (62.5)* | *1217 (65.7)* | *1816 (70.3)* |
| *Other Hispanic* | *180 (37.5)* | *635 (34.3)* | *766 (29.7)* |
| NH White | 518 (35.7) | 1191 (29.7) | 1316 (27.8) |
| Asian/Pacific Islander | 251 (17.3) | 577 (14.4) | 462 (9.8) |
| *Filipino* | *93 (37.1)* | *203 (35.2)* | *145 (31.4)* |
| *Chinese* | *44 (17.5)* | *116 (20.1)* | *91 (19.7)* |
| *Korean* | *49 (19.5)* | *119 (20.6)* | *39 (8.4)* |
| *Vietnamese* | *21 (8.4)* | *38 (6.6)* | *38 (8.2)* |
| *Japanese* | *16 (6.4)* | *35 (6.1)* | *31 (6.7)* |
| *Other Asian* | *28 (11.2)* | *66 (11.4)* | *118 (25.5)* |
| Black | 175 (12.1) | 334 (8.3) | 295 (6.2) |
| Multiracial/Other/Unknown | 19 (1.3) | 51 (1.3) | 54 (1.1) |
| American Indian/Alaskan Native | 9 (0.6) | 12 (0.3) | 22 (0.5) |
| **Educational Attainment** | | | |
| Less than High School | 345 (23.8) | 1416 (35.3) | 1876 (39.7) |
| High School | 455 (31.3) | 1188 (29.6) | 1384 (29.3) |
| Some college | 177 (12.2) | 373 (9.3) | 490 (10.4) |
| Associate Degree | 86 (5.9) | 182 (4.5) | 210 (4.4) |
| Bachelor's Degree | 199 (13.7) | 388 (9.7) | 379 (8.0) |
| Graduate Degree | 110 (7.6) | 198 (4.9) | 191 (4.0) |
| Unknown | 80 (5.5) | 272 (6.8) | 201 (4.3) |
| **Country of Birth** | | | |
| United States | 710 (48.9) | 1644 (40.9) | 2059 (43.5) |
| Mexico | 246 (16.9) | 1083 (27.0) | 1613 (34.1) |
| Philippines | 98 (6.8) | 203 (5.1) | 141 (3.0) |
| El Salvador | 46 (3.2) | 149 (3.7) | 169 (3.6) |
| Guatemala | 23 (1.6) | 104 (2.6) | 92 (1.9) |
| South Korea | 49 (3.4) | 111 (2.8) | 35 (0.7) |
| Other country | 239 (16.5) | 561 (14.0) | 502 (10.6) |
| Unknown | 41 (2.8) | 162 (4.0) | 120 (2.5) |

**eFigure 3. Age-standardized mortality rate ratios (MRR) and 95% CI by race/ethnicity and study period.**

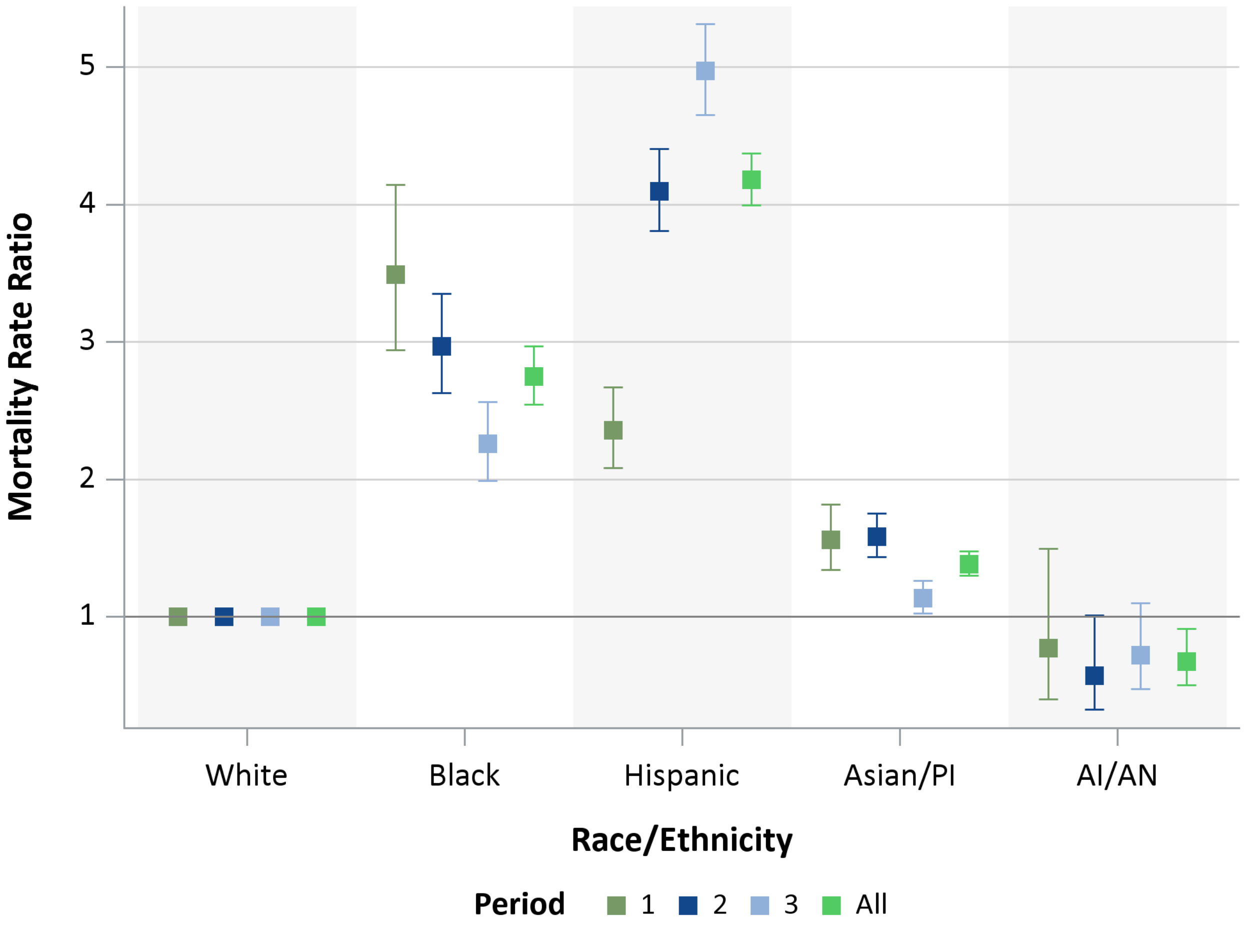


Referent group is non-Hispanic Whites.

**eTable 2. Age-standardized mortality rates (MR) and mortality rate ratios (MRR) by sex, race/ethnicity, and study period. MR are per 100,000 person-years.**

| | | Entire Study Period | | | Period 1 | | | Period 2 | | | Period 3 | | |
|---|---|---|---|---|---|---|---|---|---|---|---|---|---|
| Sex | Race/ ethnicity | COVID-19 Deaths | MR (95% CI) | MRR (95% CI) | COVID-19 Deaths | MR (95% CI) | MRR (95% CI) | COVID-19 Deaths | MR (95% CI) | MRR (95% CI) | COVID-19 Deaths | MR (95% CI) | MRR (95% CI) |
| All | White | 3025 | 27.5 (26.5-28.5) | Referent | 518 | 11.0 (10.1-12) | Referent | 1191 | 34.1 (32.2-36.0) | Referent | 1316 | 49.4 (46.8-52.1) | Referent |
| | Black | 804 | 75.4 (70.4-80.8) | 2.75 (2.54-2.97) | 175 | 38.3 (33.0-44.4) | 3.49 (2.94-4.14) | 334 | 101.1 (90.8-112.5) | 2.97 (2.63-3.35) | 295 | 111.5 (99.4-124.9) | 2.26 (1.99-2.56) |
| | Hispanic | 4914 | 114.7 (111.6-118.0) | 4.18 (3.99-4.37) | 480 | 25.9 (23.6-28.3) | 2.36 (2.08-2.67) | 1852 | 139.5 (133.3-146.0) | 4.10 (3.81-4.41) | 2582 | 245.4 (236.1-255.1) | 4.97 (4.65-5.31) |
| | Asian/PI | 1290 | 38.0 (36.0-40.1) | 1.38 (1.30-1.48) | 251 | 17.1 (15.1-19.4) | 1.56 (1.34-1.82) | 577 | 54.0 (49.7-58.5) | 1.58 (1.43-1.75) | 462 | 56.1 (51.2-61.4) | 1.14 (1.02-1.26) |
| | AI/AN | 43 | 18.6 (13.8-25.0) | 0.68 (0.50-0.91) | 9 | - | - | 12 | - | - | 22 | 35.5 (23.4-53.9) | 0.72 (0.47-1.10) |
| Female | White | 1386 | 25.0 (23.7-26.4) | Referent | 202 | 8.4 (7.3-9.7) | Referent | 582 | 33.1 (30.5-35.9) | Referent | 602 | 45.0 (41.6-48.8) | Referent |
| | Black | 373 | 68.1 (61.5-75.3) | 2.72 (2.43-3.05) | 79 | 33.9 (27.2-42.3) | 4.02 (3.10-5.22) | 161 | 94.8 (81.2-110.6) | 2.86 (2.41-3.41) | 133 | 97.0 (81.8-114.9) | 2.15 (1.78-2.60) |
| | Hispanic | 1827 | 87.7 (83.8-91.8) | 3.51 (3.27-3.76) | 175 | 19.2 (16.5-22.2) | 2.27 (1.86-2.78) | 700 | 109.0 (101.2-117.4) | 3.29 (2.95-3.68) | 952 | 185.5 (174.1-197.7) | 4.12 (3.72-4.56) |
| | Asian/PI | 632 | 36.4 (33.7-39.4) | 1.46 (1.33-1.60) | 120 | 16.0 (13.4-19.2) | 1.90 (1.52-2.38) | 289 | 53.0 (47.2-59.5) | 1.60 (1.39-1.84) | 223 | 52.7 (46.3-60.1) | 1.17 (1.00-1.37) |
| | AI/AN | 22 | 20.2 (13.3-30.7) | 0.81 (0.53-1.23) | 8 | - | - | 3 | - | - | 11 | - | - |
| Male | White | 1639 | 29.6 (28.2-31.1) | Referent | 316 | 13.4 (12.0-14.9) | Referent | 609 | 34.8 (32.1-37.6) | Referent | 714 | 53.2 (49.5-57.3) | Referent |
| | Black | 431 | 84.2 (76.6-92.6) | 2.84 (2.55-3.16) | 96 | 43.3 (35.4-52.9) | 3.24 (2.58-4.07) | 173 | 109.1 (94.0-126.6) | 3.14 (2.65-3.72) | 162 | 128.4 (110.1-149.8) | 2.41 (2.03-2.86) |
| | Hispanic | 3087 | 143.4 (138.5-148.6) | 4.84 (4.56-5.14) | 305 | 33.0 (29.5-36.9) | 2.47 (2.11-2.89) | 1152 | 172.0 (162.4-182.3) | 4.95 (4.49-5.46) | 1630 | 308.7 (294.1-324.1) | 5.80 (5.31-6.33) |
| | Asian/PI | 658 | 40.3 (37.4-43.5) | 1.36 (1.24-1.49) | 131 | 18.7 (15.8-22.2) | 1.40 (1.14-1.72) | 288 | 55.8 (49.7-62.6) | 1.61 (1.40-1.85) | 239 | 60.5 (53.3-68.6) | 1.14 (0.98-1.31) |
| | AI/AN | 21 | 16.9 (11.0-26.0) | 0.57 (0.37-0.88) | 1 | - | - | 9 | - | - | 11 | - | - |

Estimates for MR and MRR suppressed where COVID-19 deaths are less than 20.

**eFigure 4. Age-standardized mortality rate ratios (MRR) and 95% CI by race/ethnicity for selected counties.**

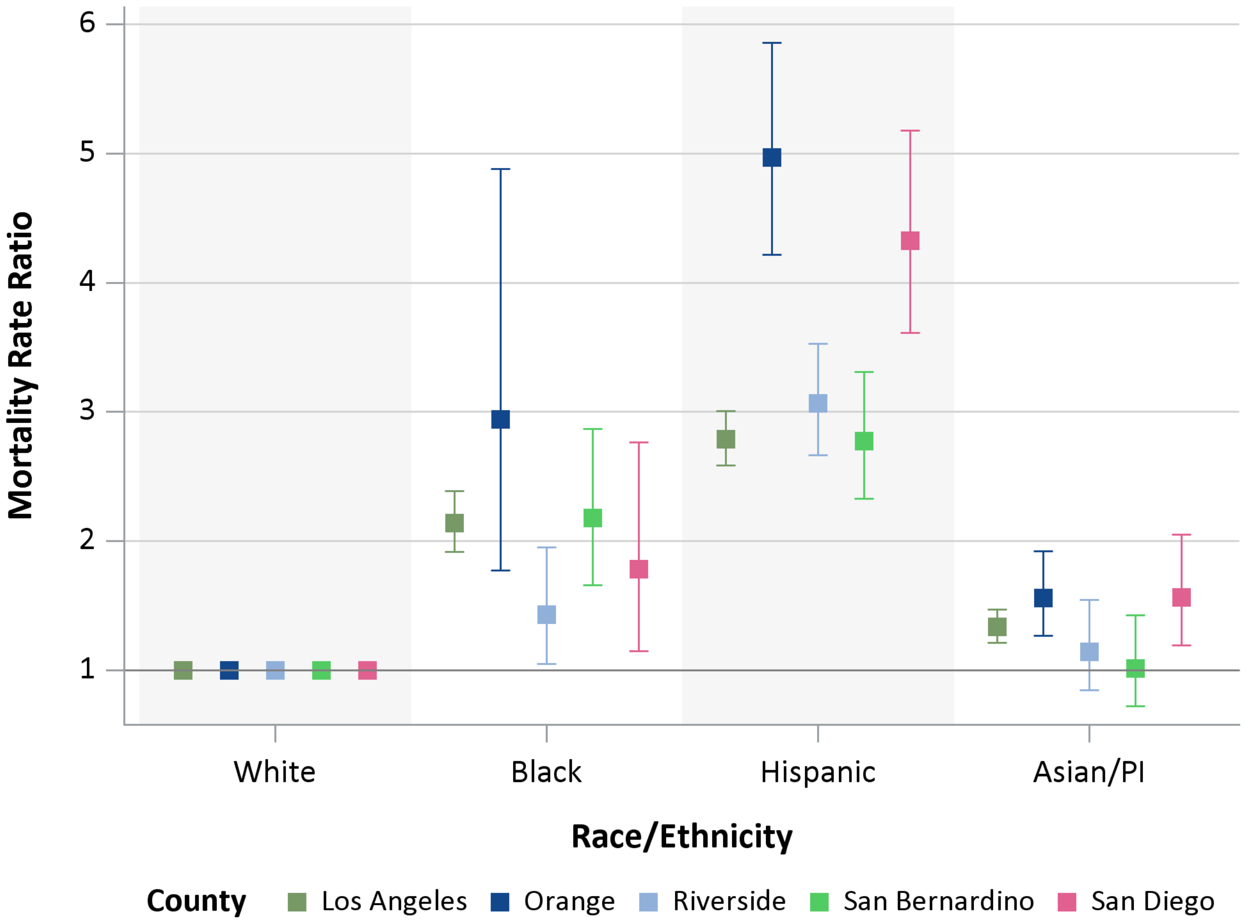


Referent group is non-Hispanic Whites.

**eTable 3. Age stratum-specific mortality rates (MR) and mortality rate ratios (MRR) by race/ethnicity and sex. MR are per 100,000 person-years.**

| | | All | | | Females | | | Males | | |
|---|---|---|---|---|---|---|---|---|---|---|
| **Race/ ethnicity** | **Age** | **COVID-19 Deaths** | **MR (95% CI)** | **MRR (95% CI)** | **COVID-19 Deaths** | **MR (95% CI)** | **MRR (95% CI)** | **COVID-19 Deaths** | **MR (95% CI)** | **MRR (95% CI)** |
| White | 20 to 54 | 104 | 2.9 (2.4-3.6) | Referent | 30 | 1.8 (1.2-2.5) | Referent | 74 | 4.0 (3.2-5.1) | Referent |
| | 55 to 64 | 242 | 21.0 (18.5-23.8) | Referent | 81 | 14.0 (11.3-17.4) | Referent | 161 | 28.0 (24.0-32.7) | Referent |
| | 65 to 74 | 557 | 59.3 (54.6-64.4) | Referent | 184 | 37.4 (32.4-43.2) | Referent | 373 | 83.2 (75.1-92.0) | Referent |
| | 75 to 84 | 885 | 181.1 (169.6-193.5) | Referent | 385 | 143.3 (129.7-158.4) | Referent | 500 | 227.2 (208.1-248.0) | Referent |
| | 85+ | 1236 | 558.5 (528.2-590.5) | Referent | 705 | 512.5 (476.0-551.7) | Referent | 531 | 634.1 (582.4-690.4) | Referent |
| Black | 20 to 54 | 80 | 13.8 (11.1-17.1) | 4.68 (3.50-6.26) | 27 | 9.5 (6.5-13.8) | 5.37 (3.19-9.04) | 53 | 17.8 (13.6-23.4) | 4.43 (3.11-6.30) |
| | 55 to 64 | 126 | 82.3 (69.1-98) | 3.92 (3.16-4.86) | 47 | 59.2 (44.4-78.7) | 4.23 (2.95-6.06) | 79 | 107.2 (86.0-133.6) | 3.82 (2.92-5.00) |
| | 65 to 74 | 196 | 211.1 (183.5-242.8) | 3.56 (3.03-4.19) | 73 | 142.0 (112.9-178.6) | 3.80 (2.89-4.98) | 123 | 297.3 (249.2-354.8) | 3.58 (2.92-4.38) |
| | 75 to 84 | 219 | 480.5 (420.9-548.5) | 2.65 (2.29-3.08) | 117 | 433.5 (361.7-519.7) | 3.02 (2.46-3.72) | 102 | 548.7 (451.9-666.2) | 2.41 (1.95-2.99) |
| | 85+ | 183 | 1057.9 (915.2-1222.8) | 1.89 (1.62-2.21) | 109 | 951.7 (788.8-1148.2) | 1.86 (1.52-2.27) | 74 | 1265.8 (1007.9-1589.7) | 2.00 (1.57-2.54) |
| Hispanic | 20 to 54 | 854 | 24.9 (23.3-26.7) | 8.48 (6.92-10.39) | 245 | 14.7 (13.0-16.7) | 8.34 (5.71-12.18) | 609 | 34.6 (32.0-37.5) | 8.58 (6.74-10.92) |
| | 55 to 64 | 954 | 152.4 (143.0-162.4) | 7.26 (6.30-8.36) | 260 | 81.2 (71.9-91.7) | 5.81 (4.52-7.45) | 694 | 227.1 (210.8-244.7) | 8.10 (6.82-9.62) |
| | 65 to 74 | 1190 | 349.7 (330.4-370.1) | 5.90 (5.33-6.52) | 391 | 211.1 (191.2-233.1) | 5.64 (4.74-6.72) | 799 | 514.6 (480.1-551.5) | 6.19 (5.47-7.00) |
| | 75 to 84 | 1001 | 636.9 (598.7-677.6) | 3.52 (3.21-3.85) | 429 | 466.7 (424.5-513.0) | 3.26 (2.84-3.74) | 572 | 877.4 (808.4-952.4) | 3.86 (3.43-4.35) |
| | 85+ | 912 | 1416.5 (1327.5-1511.5) | 2.54 (2.33-2.76) | 502 | 1229.4 (1126.4-1341.7) | 2.40 (2.14-2.69) | 410 | 1741.0 (1580.4-1918.0) | 2.75 (2.41-3.12) |
| Asian/PI | 20 to 54 | 78 | 4.5 (3.6-5.7) | 1.54 (1.15-2.06) | 24 | 2.6 (1.8-3.9) | 1.50 (0.88-2.57) | 54 | 6.6 (5.1-8.6) | 1.64 (1.16-2.33) |
| | 55 to 64 | 125 | 31.2 (26.1-37.1) | 1.48 (1.20-1.84) | 37 | 16.9 (12.2-23.3) | 1.21 (0.82-1.78) | 88 | 48.3 (39.2-59.6) | 1.72 (1.33-2.23) |
| | 65 to 74 | 228 | 79.5 (69.8-90.5) | 1.34 (1.15-1.56) | 84 | 52.5 (42.4-65.1) | 1.40 (1.08-1.82) | 144 | 113.8 (96.7-134.0) | 1.37 (1.13-1.66) |
| | 75 to 84 | 302 | 211.5 (188.9-236.7) | 1.17 (1.02-1.33) | 130 | 159.7 (134.5-189.7) | 1.11 (0.91-1.36) | 172 | 279.8 (240.9-324.9) | 1.23 (1.04-1.46) |
| | 85+ | 556 | 848.5 (780.8-922) | 1.52 (1.37-1.68) | 356 | 870.1 (784.2-965.3) | 1.70 (1.50-1.93) | 200 | 812.6 (707.4-933.4) | 1.28 (1.09-1.51) |

Results for the Native American/Alaskan Native group not shown due to small numbers.

**eTable 4. Proportionate mortality (PM) and proportionate mortality ratio (PMR) by selected characteristics for COVID-19 deaths occurring between March 1 - July 31, 2020 among decedents aged 20 years and older.**

| | COVID-19 Deaths | PM (95% CI)* | PMR (95% CI) |
|---|---|---|---|
| **Sex** | | | |
| Female | 4283 | 8.2 (8.0-8.4) | Referent |
| Male | 5910 | 10.4 (10.2-10.7) | 1.27 (1.22-1.32) |
| **Race/Ethnicity** | | | |
| White | 3024 | 4.5 (4.4-4.7) | Referent |
| Black | 804 | 9.7 (9.0-10.4) | 2.14 (1.98-2.30) |
| Hispanic | 4910 | 23.8 (23.1-24.5) | 5.25 (5.03-5.48) |
| Asian/PI | 1288 | 11.5 (10.9-12.1) | 2.53 (2.38-2.70) |
| American Indian/Alaskan Native | 43 | 6.7 (5.0-9.1) | 1.49 (1.11-1.99) |
| **Educational Attainment** | | | |
| Less than High School | 3635 | 17.9 (17.3-18.5) | 2.86 (2.70-3.03) |
| High School | 3025 | 8.0 (7.7-8.3) | 1.28 (1.20-1.36) |
| Some college or Associate Degree | 1516 | 6.2 (5.9-6.5) | 0.99 (0.92-1.06) |
| Bachelor or Graduate Degree | 1464 | 6.3 (5.9-6.6) | Referent |
| **Ethnicity and Nativity Combined** | | | |
| U.S.-born non-Hispanic | 3271 | 4.7 (4.5-4.9) | Referent |
| U.S.-born Hispanic | 1137 | 12.0 (11.4-12.8) | 2.56 (2.40-2.72) |
| Foreign-born non-Hispanic | 1753 | 10.2 (9.7-10.6) | 2.16 (2.04-2.28) |
| Foreign-born Hispanic | 3709 | 33.6 (32.5-34.7) | 7.14 (6.84-7.45) |

*Sub-population PM were calculated using all-cause mortality in the same sub-population occurring March 1-July 31 in 2016-2019 among decedents aged 20 years and older.

**eTable 5. Proportionate mortality (PM) and proportionate mortality ratio (PMR) by race/ethnicity and age for COVID-19 deaths occurring between March 1 - July 31, 2020 among decedents aged 20 years and older.**

| Race/ethnicity | Age | COVID-19 Deaths | PM (95% CI)* | PMR (95% CI) |
|---|---|---|---|---|
| White | 20 to 54 | 104 | 2.0 (1.6-2.4) | Referent |
| | 55 to 64 | 242 | 3.1 (2.8-3.5) | Referent |
| | 65 to 74 | 557 | 4.6 (4.2-5.0) | Referent |
| | 75 to 84 | 885 | 5.5 (5.2-5.9) | Referent |
| | 85+ | 1236 | 4.8 (4.6-5.1) | Referent |
| Black | 20 to 54 | 80 | 5.4 (4.3-6.7) | 2.76 (2.07-3.67) |
| | 55 to 64 | 126 | 7.7 (6.4-9.1) | 2.45 (1.99-3.02) |
| | 65 to 74 | 196 | 10.7 (9.3-12.3) | 2.34 (2.00-2.73) |
| | 75 to 84 | 219 | 12.6 (11.0-14.3) | 2.27 (1.97-2.61) |
| | 85+ | 183 | 11.4 (9.9-13.2) | 2.37 (2.04-2.74) |
| Hispanic | 20 to 54 | 854 | 18.9 (17.6-20.2) | 9.62 (7.88-11.74) |
| | 55 to 64 | 953 | 29.3 (27.5-31.2) | 9.38 (8.20-10.74) |
| | 65 to 74 | 1190 | 32.9 (31.1-34.9) | 7.20 (6.56-7.91) |
| | 75 to 84 | 1001 | 24.0 (22.6-25.6) | 4.34 (3.99-4.72) |
| | 85+ | 912 | 17.9 (16.8-19.1) | 3.71 (3.42-4.02) |
| Asian/Pacific Islander | 20 to 54 | 78 | 6.9 (5.5-8.6) | 3.53 (2.65-4.70) |
| | 55 to 64 | 125 | 9.8 (8.2-11.7) | 3.15 (2.56-3.87) |
| | 65 to 74 | 227 | 11.9 (10.4-13.5) | 2.60 (2.24-3.01) |
| | 75 to 84 | 302 | 10.9 (9.7-12.2) | 1.96 (1.73-2.22) |
| | 85+ | 556 | 13.4 (12.4-14.6) | 2.78 (2.53-3.05) |

*Sub-population PM were calculated using all-cause mortality in the same sub-population occurring March 1-July 31 in 2016-2019 among decedents aged 20 years and older.

**eTable 6. Proportionate mortality (PM) and proportionate mortality ratio (PMR) by race/ethnicity, age, and educational attainment for COVID-19 deaths occurring between March 1 - July 31, 2020 among decedents aged 20 years and older.**

| | | High school or less | | | Some college or more | | |
|---|---|---|---|---|---|---|---|
| **Race/ethnicity** | **Age** | **COVID-19 Deaths** | **PM (95% CI)*** | **PMR (95% CI)** | **COVID-19 Deaths** | **PM (95% CI)*** | **PMR (95% CI)** |
| White | 20 to 64 | 191 | 3.0 (2.6-3.4) | Referent | 134 | 2.2 (1.9-2.6) | Referent |
| | 65 to 74 | 238 | 4.9 (4.3-5.6) | Referent | 268 | 3.9 (3.5-4.4) | Referent |
| | 75 to 84 | 407 | 5.7 (5.2-6.3) | Referent | 426 | 5.0 (4.5-5.5) | Referent |
| | 85+ | 645 | 5.1 (4.7-5.5) | Referent | 535 | 4.3 (3.9-4.6) | Referent |
| Black | 20 to 64 | 115 | 6.9 (5.7-8.3) | 2.32 (1.86-2.91) | 83 | 6.4 (5.1-7.9) | 2.88 (2.21-3.77) |
| | 65 to 74 | 101 | 11.9 (9.8-14.5) | 2.43 (1.94-3.03) | 74 | 8.4 (6.7-10.5) | 2.14 (1.67-2.75) |
| | 75 to 84 | 112 | 12.5 (10.4-15.1) | 2.18 (1.79-2.66) | 91 | 11.4 (9.3-14.0) | 2.28 (1.84-2.83) |
| | 85+ | 112 | 11.8 (9.8-14.2) | 2.32 (1.92-2.81) | 61 | 10.1 (7.9-13.0) | 2.37 (1.84-3.05) |
| Hispanic | 20 to 64 | 1452 | 25.3 (24.0-26.7) | 8.54 (7.37-9.88) | 292 | 15.5 (13.9-17.4) | 7.03 (5.76-8.56) |
| | 65 to 74 | 966 | 35.8 (33.6-38.1) | 7.30 (6.39-8.35) | 162 | 19.9 (17.0-23.2) | 5.09 (4.24-6.09) |
| | 75 to 84 | 858 | 25.8 (24.1-27.6) | 4.49 (4.02-5.02) | 99 | 13.3 (10.9-16.2) | 2.68 (2.18-3.29) |
| | 85+ | 785 | 18.4 (17.2-19.8) | 3.63 (3.29-4.01) | 83 | 11.6 (9.4-14.4) | 2.72 (2.19-3.39) |
| Asian/Pacific Islander | 20 to 64 | 64 | 6.6 (5.1-8.4) | 2.22 (1.68-2.92) | 134 | 9.7 (8.2-11.5) | 4.38 (3.48-5.53) |
| | 65 to 74 | 72 | 9.1 (7.2-11.5) | 1.85 (1.44-2.39) | 145 | 13.4 (11.4-15.8) | 3.43 (2.83-4.15) |
| | 75 to 84 | 140 | 10.4 (8.8-12.3) | 1.81 (1.51-2.18) | 150 | 10.9 (9.3-12.8) | 2.19 (1.83-2.61) |
| | 85+ | 344 | 13.4 (12.1-14.9) | 2.65 (2.34-3.00) | 198 | 13.1 (11.4-15.0) | 3.07 (2.63-3.58) |

*Sub-population PM were calculated using all-cause mortality in the same sub-population occurring March 1-July 31 in 2016-2019 among decedents aged 20 years and older.

**eFigure 5. Crude all-cause mortality rate ratios and 95% CI based on 2019 mortality data and population estimates by race/ethnicity and age.**

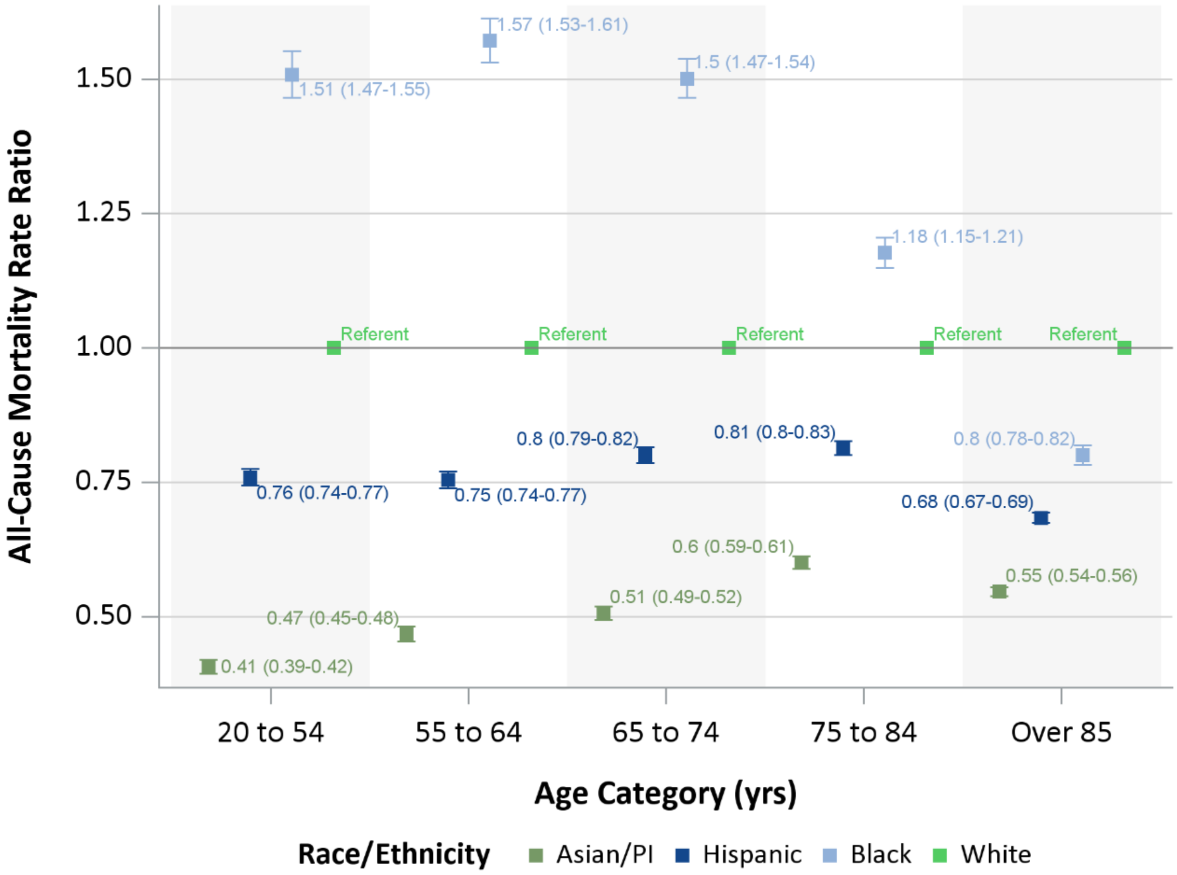


Referent group is non-Hispanic Whites.

| eTable 7. Crude all-cause mortality rate ratios for 2019, proportionate mortality ratio (PMR) for COVID-19, and adjusted PMR by race/ethnicity and age. The PMR are for COVID-19 deaths occurring between March 1 - July 31, 2020 among decedents aged 20 years and older. | | | | |
|---|---|---|---|---|
| **Race/ethnicity** | **Age** | **2019 all-cause mortality rate ratio** | **PMR** | **Adjusted* PMR** |
| White | 20 to 54 | 1.00 | 1.00 | 1.00 |
| | 55 to 64 | 1.00 | 1.00 | 1.00 |
| | 65 to 74 | 1.00 | 1.00 | 1.00 |
| | 75 to 84 | 1.00 | 1.00 | 1.00 |
| | 85<= | 1.00 | 1.00 | 1.00 |
| Black | 20 to 54 | 1.51 | 2.76 | 4.16 |
| | 55 to 64 | 1.57 | 2.45 | 3.85 |
| | 65 to 74 | 1.50 | 2.34 | 3.51 |
| | 75 to 84 | 1.18 | 2.27 | 2.67 |
| | 85<= | 0.80 | 2.37 | 1.89 |
| Hispanic | 20 to 54 | 0.76 | 9.62 | 7.30 |
| | 55 to 64 | 0.75 | 9.38 | 7.08 |
| | 65 to 74 | 0.80 | 7.20 | 5.76 |
| | 75 to 84 | 0.81 | 4.34 | 3.53 |
| | 85<= | 0.68 | 3.71 | 2.54 |
| Asian/Pacific Islander | 20 to 54 | 0.41 | 3.53 | 1.44 |
| | 55 to 64 | 0.47 | 3.15 | 1.47 |
| | 65 to 74 | 0.51 | 2.60 | 1.31 |
| | 75 to 84 | 0.60 | 1.96 | 1.18 |
| | 85<= | 0.55 | 2.78 | 1.52 |
| *PMR were indirectly adjusted for differences in 2019 all-cause mortality by multiplying PMR by the stratum specific all-cause mortality rate ratio. | | | | |

**eTable 8. Proportionate mortality (PM) and proportionate mortality ratio (PMR) by ethnicity and nativity combined and age for COVID-19 deaths occurring between March 1 - July 31, 2020 among decedents aged 20 years and older.**

| Ethnicity and Nativity Combined | Age | COVID-19 Deaths | PM (95% CI)* | PMR (95% CI) |
|---|---|---|---|---|
| U.S.-born non-Hispanic | 20 to 54 | 182 | 2.6 (2.3-3.1) | Referent |
| | 55 to 64 | 346 | 3.9 (3.5-4.3) | Referent |
| | 65 to 74 | 646 | 4.9 (4.6-5.3) | Referent |
| | 75 to 84 | 931 | 5.8 (5.4-6.2) | Referent |
| | 85<= | 1166 | 4.8 (4.5-5.0) | Referent |
| U.S.-born Hispanic | 20 to 54 | 247 | 9.7 (8.6-11.0) | 3.67 (3.04-4.42) |
| | 55 to 64 | 160 | 11.2 (9.6-13.1) | 2.89 (2.42-3.46) |
| | 65 to 74 | 243 | 16.5 (14.6-18.7) | 3.35 (2.92-3.85) |
| | 75 to 84 | 211 | 12.9 (11.2-14.7) | 2.22 (1.93-2.55) |
| | 85<= | 276 | 11.7 (10.4-13.1) | 2.45 (2.17-2.78) |
| Foreign-born non-Hispanic | 20 to 54 | 88 | 6.9 (5.6-8.5) | 2.62 (2.04-3.35) |
| | 55 to 64 | 137 | 7.7 (6.5-9.1) | 1.98 (1.64-2.40) |
| | 65 to 74 | 296 | 10.6 (9.5-11.9) | 2.15 (1.89-2.46) |
| | 75 to 84 | 443 | 9.9 (9.0-10.8) | 1.70 (1.53-1.90) |
| | 85<= | 789 | 11.4 (10.6-12.2) | 2.40 (2.20-2.61) |
| Foreign-born Hispanic | 20 to 54 | 597 | 30.6 (28.3-33.2) | 11.59 (9.89-13.57) |
| | 55 to 64 | 782 | 43.6 (40.7-46.8) | 11.25 (10.02-12.64) |
| | 65 to 74 | 924 | 43.9 (41.2-46.8) | 8.91 (8.14-9.74) |
| | 75 to 84 | 778 | 31.2 (29.1-33.5) | 5.39 (4.95-5.87) |
| | 85<= | 628 | 23.3 (21.5-25.1) | 4.89 (4.48-5.34) |

*Sub-population PM were calculated using all-cause mortality in the same sub-population occurring March 1-July 31 in 2016-2019 among decedents aged 20 years and older.
